# The Association of QRS Duration with Risk of Adverse Outcomes in Sex- and Race- Based Subgroups: The Dallas Heart Study

**DOI:** 10.1101/2023.05.15.23290016

**Authors:** Nitin Kondamudi, Yihun Zeleke, Anna Rosenblatt, Gene Hu, Christopher Grubb, Mark S. Link

**Affiliations:** Division of Cardiology, Department of Internal Medicine, University of Texas Southwestern Medical Center, 5323 Harry Hines Blvd, Dallas, TX

## Abstract

**Introduction:** We explored sex and race differences in the prognostic implications of QRS prolongation among healthy adults.

**Methods:** Participants from the Dallas Heart Study (DHS) free of cardiovascular (CV) disease who underwent ECG testing and cMRI evaluation were included. Multivariable linear regression was used to examine the cross-sectional association of QRS duration with left ventricular (LV) mass, LV ejection fraction (LVEF), and LV end diastolic volume (LVEDV). Association of QRS duration with risk of MACE was evaluated using Cox models. Interaction testing was performed between QRS duration and sex/race respectively for each outcome of interest. QRS duration was log transformed.

**Results:** The study included 2,785 participants. Longer QRS duration was associated with higher LV mass, lower LVEF, and higher LVEDV, independent of CV risk factors ([β: 0.21, P<0.001], [β: - 0.13, P<0.001], [β: 0.22, P<0.001] respectively). Men with longer QRS duration were more likely to have higher LV mass and higher LVEDV compared to women (P-int=0.012, P-int=0.01, respectively). Black participants with longer QRS duration were more likely to have higher LV mass as compared to White participants (P-int<0.001). In Cox analysis, QRS prolongation was associated with higher risk of MACE in women (HR = 6.66 [95% CI: 2.32, 19.1]) but not men. This association was attenuated after adjustment for CV risk factors, with a trend toward significance (HR = 2.45 [95% CI: 0.94, 6.39]). Longer QRS duration was not associated with risk of MACE in Black or White participants in the adjusted models. No interaction between sex/race and QRS duration for risk of MACE was observed.

**Discussion:** In healthy adults, QRS duration is differentially associated with abnormalities in LV structure and function. These findings inform the use of QRS duration in identifying subgroups at risk for CV disease, and caution against using QRS duration cut offs uniformly for clinical decision making.

**What is known?:** QRS prolongation in healthy adults is associated with higher risk of death, cardiovascular disease, and left ventricular hypertrophy.

**What the study adds?:** QRS prolongation may reflect a higher degree of underlying LV hypertrophy in Blacks compared to Whites. Longer QRS interval may reflect higher risk of adverse cardiac events, driven by prevalent cardiovascular risk factors.

**Graphic Abstract:** Risk of underlying left ventricular hypertrophy in demographic groups based on QRS prolongation

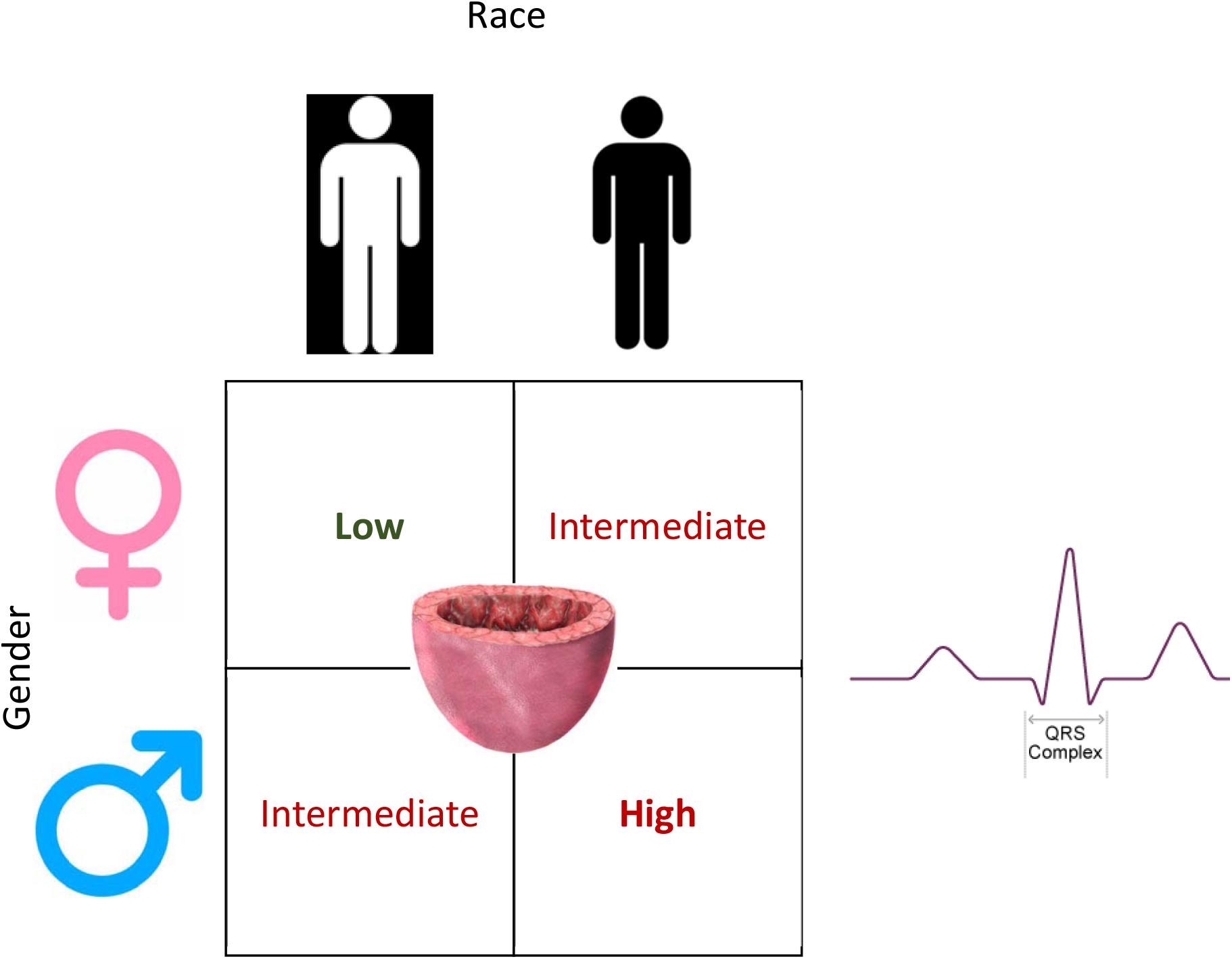

## Introduction

The QRS complex on a 12-lead electrocardiogram represents the electrical depolarization of the ventricular myocardium. Prolongation in QRS duration is associated with higher risk of adverse outcomes among individuals with cardiovascular disease (CVD)(1, 2, 3, 4, 5, 6, 7, 8, 9). In adults with heart failure with reduced ejection fraction (HFrEF), QRS duration and morphology are used to identify patients who may benefit from cardiac resynchronization therapy (CRT)(11). Notably, differential improvement in clinical outcomes for CRT has been observed, such that women with left bundle branch block (LBBB) and QRS duration > 150 ms derive the most benefit(12).

In populations free of CVD, studies have shown that prolonged QRS duration is associated with greater risk of all cause death, CV mortality, and sudden cardiac death (13, 14, 15, 16). However, sex- and race-based differences in the prognostic implications of QRS duration in healthy adults are not well known. Moreover, the mechanisms that underlie the association of QRS duration with risk of adverse events in these subgroups remains unclear. The purpose of this study was to examine whether sex or race modified the association of QRS duration with left ventricular structure and function, and subsequent risk of major adverse cardiac events (MACE) in a multiethnic healthy cohort. We hypothesized that QRS duration would be associated with LV hypertrophy, LV systolic dysfunction, and risk of adverse outcomes in sex- and race-based subgroups.

## Methods

### Study Population

The Dallas Heart Study (DHS) is a multiethnic, probability-based, population cohort study of Dallas County residents with a deliberate oversampling of self-identified Black individuals. The methods and study design of DHS have been described previously(17). Briefly, the DHS was conducted over three visits at the University of Texas Southwestern Medical Center. Individuals enrolled in DHS-1 underwent initial examination from 2000 to 2002 with cardiovascular risk factor evaluation, laboratory testing, and imaging studies. All study participants provided written informed consent. The University of Texas Southwestern Medical Center Institutional Review Board approved the protocol for DHS-1. DHS phase-1 enrolled 6,101 participants, of whom 763 were excluded due to prevalent CVD. Of the remaining participants, 3,066 participants underwent both ECG and cardiac magnetic resonance imaging (cMRI) evaluation. Participants missing data were excluded (N=281). No participants in the cohort had a paced rhythm. The final study sample included 2,785 participants (**Figure 1**).

**Figure 1:**
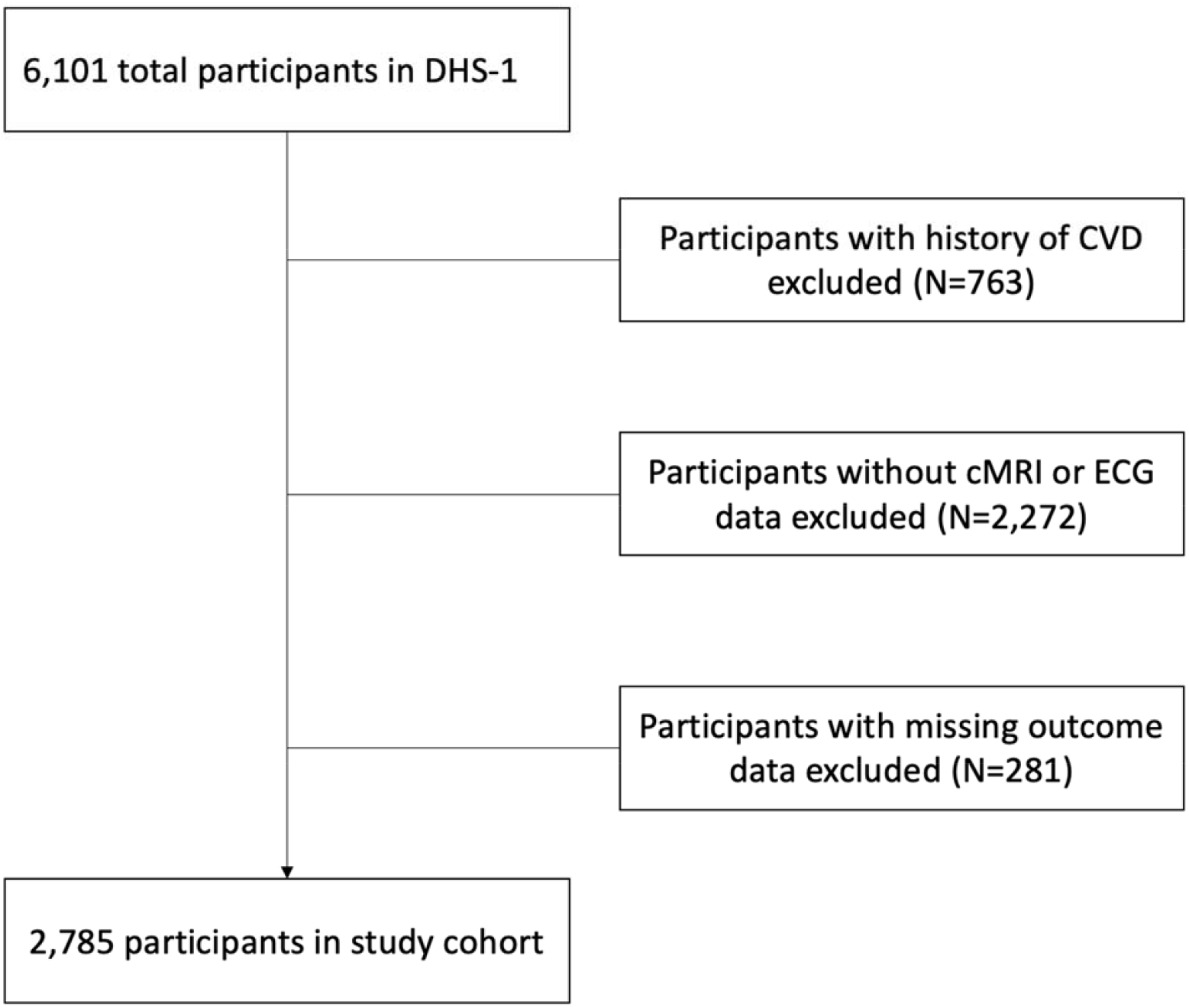
Study consort diagram Abbreviations- DHS-1: Dallas heart study 1, CVD: cardiovascular disease, cMRI: cardiac magnetic resonance imaging

### ECG

ECGs measurements including PR and QT intervals, QRS duration, J-point elevation, and heart rate were calculated by the automated Marquette 12-SL analysis system (General Electric, Boston, Massachusetts)(18). Right bundle branch block (RBBB), LBBB, and non-specific intraventricular conduction delay (NIVCD) were described according to the 2018 Heart Rhythm Society/American College of Cardiology/American Heart Association (ACC/AHA/HRS) guidelines. RBBB was defined as typical RBBB morphology with a QRS duration ≥ 120 ms. LBBB was defined as typical LBBB morphology with a QRS duration ≥ 120 ms. NIVCD was defined by QRS > 110 ms, without morphological designation of RBBB or LBBB (19). Automated ECG reads were reviewed by a board-certified cardiologist.

### cMRI

CMR was performed using 2 comparable 1.5 T systems (Phillips Medical Systems, Best, The Netherlands). The DHS-1 CMR protocol has been previously described.(20) Measurements of mass and volume were obtained from short-axis breath-hold electrocardiography-gated cine CMR using MASS software (Medis Medical Imaging Systems, Leiden, the Netherlands)(21). Epicardial and endocardial borders were contoured at LV end-diastole. LV mass was calculated using the product of the difference in epicardial and endocardial areas multiplied by slice thickness and section gap multiplied by the specific gravity of the myocardium. To calculate LV mass index, LV mass was divided by body surface area. Stroke volume was calculated by LV end-diastolic volume minus LV end-systolic volume. The EF was estimated by stroke volume divided by the LV end-diastolic volume X 100%.

### Outcomes

The primary outcome was a composite of non-fatal myocardial infarction, non-fatal cerebrovascular accident, surgical or percutaneous revascularization, heart failure hospitalization, and all-cause death (MACE). Non-fatal CV events were adjudicated by a blinded endpoint committee using a detailed health survey for interval CV events and quarterly assessment of admissions from the Dallas-Fort Worth Hospital Council Data Initiative Database (22). The national death index was used to adjudicate death events. All outcome events were adjudicated through 2016.

### Covariates

Assessment of demographic, clinical, and anthropometric characteristics in DHS have been previously described(17). Race/ethnicity and sex were self-reported using questionnaires. Height, weight, and body surface area were measured by standard scales(23). Body mass index (BMI) was computed by dividing weight in kilograms by height in meters squared. Hypertension was defined as having blood pressure >140/90 mmHg or taking antihypertensive medication(s) at the time of enrollment. Diabetes mellitus was defined as having fasting glucose >126 mg/dL or taking hypoglycemic medications. Hypercholesterolemia was defined as low density lipoprotein cholesterol > 160 mg/dL on a fasting sample, direct low-density lipoprotein >160 mg/dL on non-fasting sample, total cholesterol >240 mg/dL, or treatment with a statin medication(24). Smoking history was defined as smoking > 100 lifetime cigarettes and/or smoking within the last 30 days.

### Statistical Analysis

The study population was stratified into quartiles of QRS duration. Clinical characteristics across quartiles of QRS duration were compared using Kruskal-Wallis rank test for continuous variables and Chi-Square test for categorical variables. Continuous variables were reported as median with interquartile range (or mean with standard deviation if the data was not normally distributed). Categorical variables were reported as percentage. The distribution of QRS duration in men vs women and White vs Black participants were displayed using violin plots and compared using Mann-Whitney testing respectively.

Multivariable linear regression models were used to examine the cross-sectional association of QRS duration (independent variable) as a continuous measure with LV mass index, LV end diastolic volume (LVEDV) index, and LVEF (dependent variables), respectively. Separate multivariable adjusted linear regression models were constructed for each dependent variable with adjustment for age, race, ethnicity, gender, smoking status, BMI, prevalent hypertension, prevalent diabetes, systolic blood pressure, heart rate, and serum fasting glucose.

The unadjusted association of QRS duration by quartile with risk of MACE was assessed using Kaplan Meier estimates and log rank testing. Cox proportional hazard models were used to examine the association of QRS duration as a continuous measure with risk of MACE in the total study population. The model was adjusted for age, race, ethnicity, gender, smoking status, BMI, prevalent hypertension, prevalent diabetes, systolic blood pressure, heart rate, and serum fasting glucose. The analysis was replicated in subgroups stratified by sex and race. Given the distribution of QRS duration was skewed in our study population, QRS duration was log transformed for the analysis. Statistical analysis was performed using R.

## Results

The final study population for the survival analysis included 2,785 participants (32.4% White, 49.8% Black, mean age 44.1 years). Over a median follow-up time of 14.2 years the primary outcome event occurred in 423 participants. Baseline characterization of the study population are displayed in **Table 1**. QRS duration ranged from 60 msec to 164 msec. Participants with prolonged QRS duration were more likely to be older, White, male, and had higher systolic blood pressure, higher serum high sensitivity (hs) troponin, lower circulating NT-ProBNP, and lower triglyceride levels. In terms of indices of LV structure and function, participants with prolonged QRS duration had higher LV mass index, lower LVEF, and higher LVEDV index.

**Table 1:**
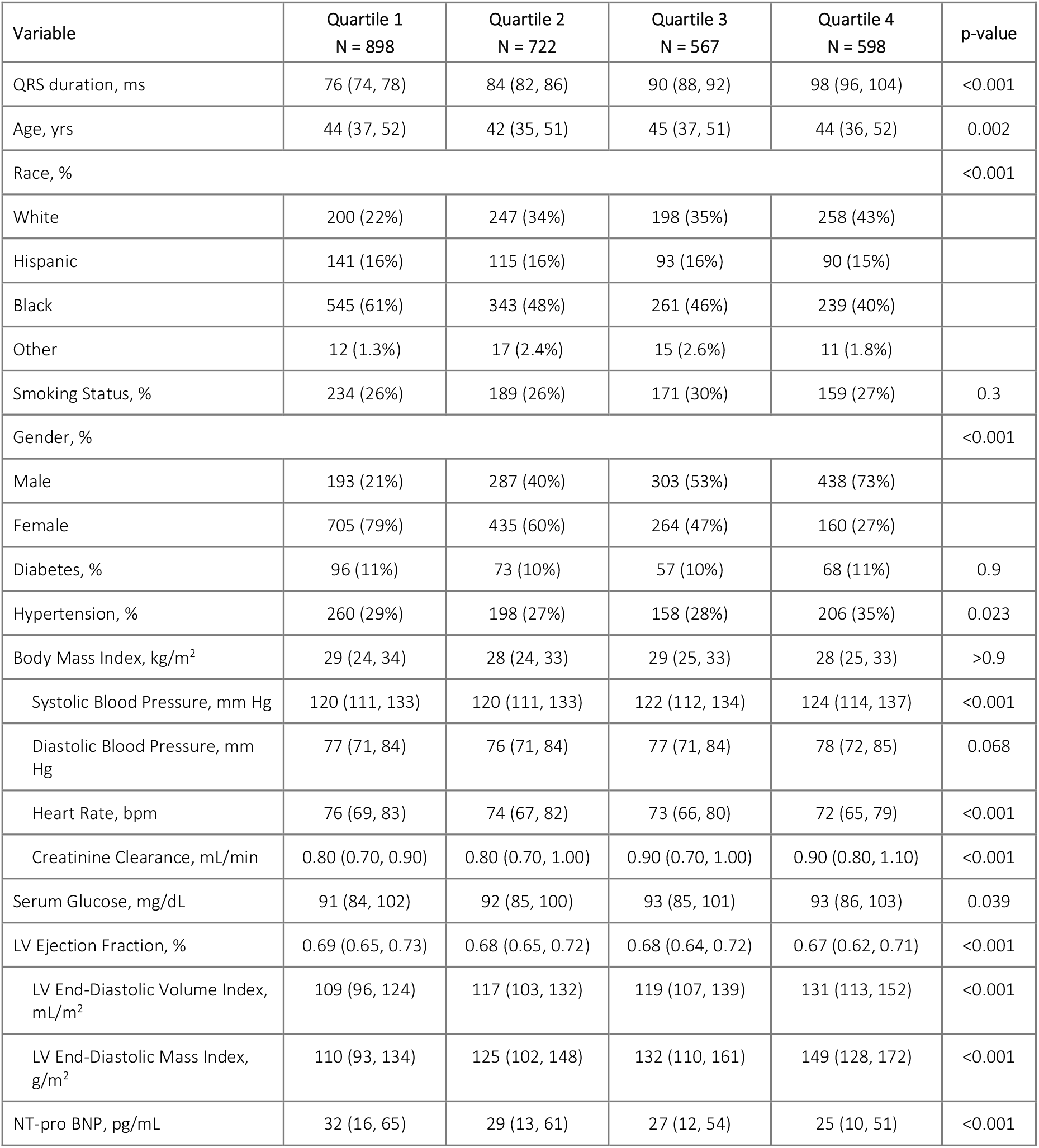
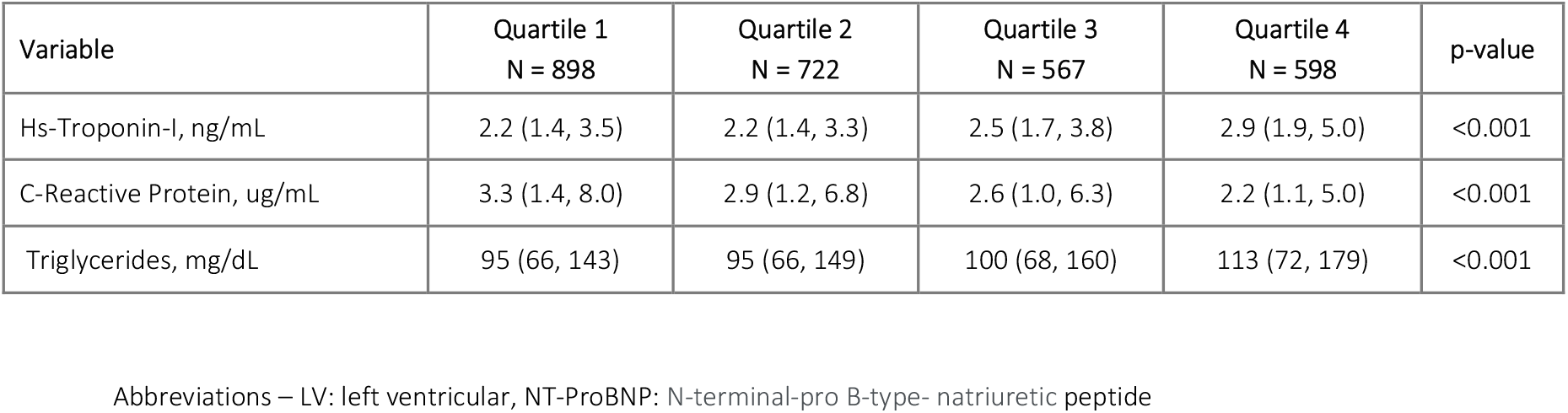
Characteristics of the Study Population Stratified by Quartiles of QRS Duration

Prevalent hypertension, diabetes, smoking status, and BMI did not differ significantly across quartiles of QRS duration. Black participants had shorter QRS duration as compared to White participants (P<0.001), and women had shorter QRS duration as compared to men (P<0.001). Box whisker plots comparing the distribution of QRS duration in Black vs White participants and men vs women are displayed in **Figure 2**.

**Figure 2:**
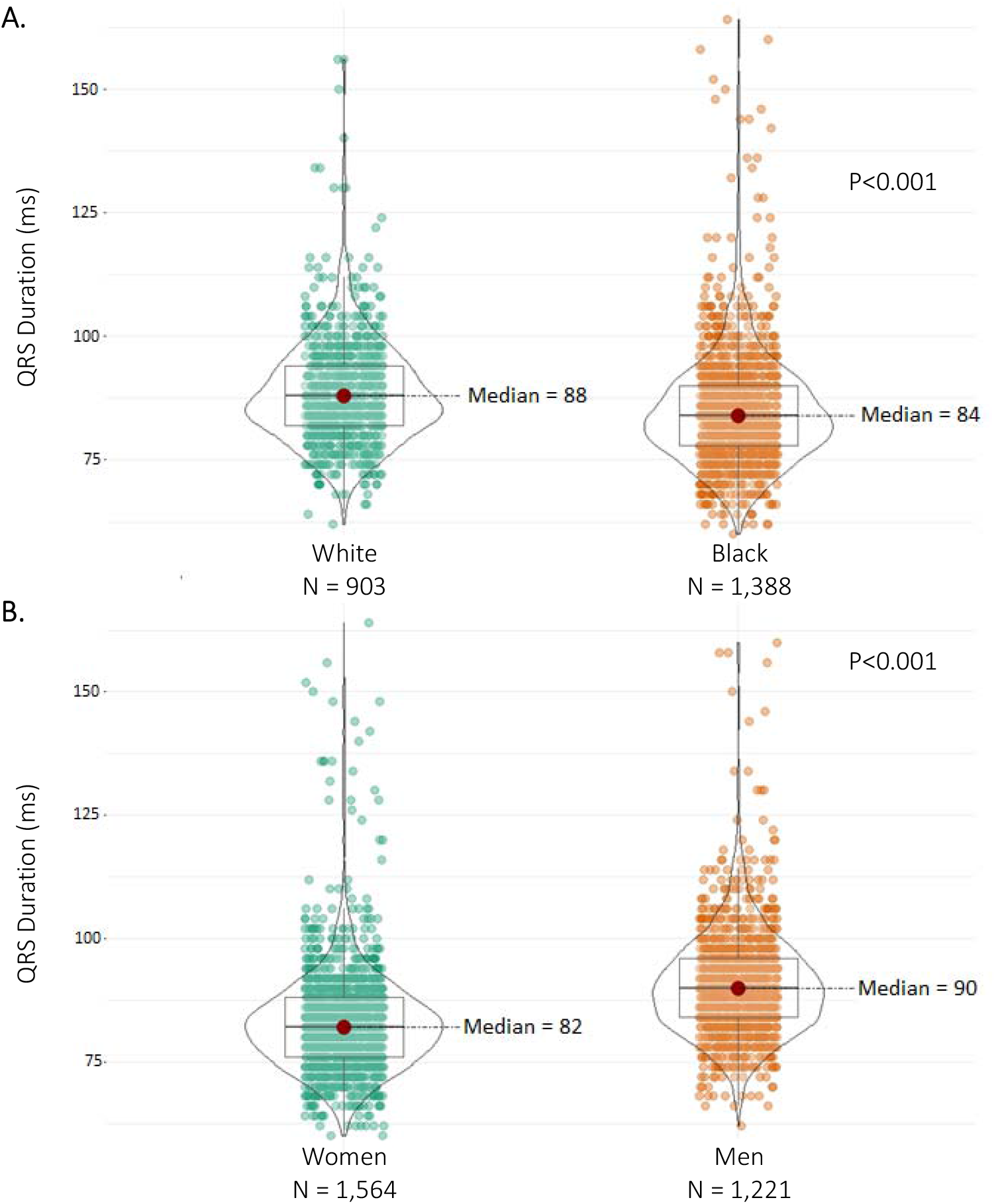
Distribution of QRS duration in Black vs White participants (A.) and men vs women (B.) Mann-Whitney test was used to compare the distribution of QRS duration in Black vs White participants and men vs women respectively.

In multivariable linear regression analysis, longer QRS duration was independently associated with higher LV mass index (β: 0.21, P<0.001), lower LVEF (β: -0.13, P<0.001), and higher LVEDV index (β: 0.22, P<0.001) after adjustment for CV risk factors. This pattern of association was consistent across sex- and race-based subgroups with significant interaction noted between sex/race, QRS duration, and indices of LV structure and function (**Table 2**).

**Table 2:** Multivariable Adjusted Cross-sectional Association of log transformed QRS duration with LV Structure and Function

|  | Total |  | Black |  | White |  | P*Int | Men |  | Women |  | P*Int |
| --- | --- | --- | --- | --- | --- | --- | --- | --- | --- | --- | --- | --- |
| Imaging parameter | SE ($\beta$) | p-value | SE ($\beta$) | p-value | SE ($\beta$) | p-value | | SE ($\beta$) | p-value | SE ($\beta$) | p-value | |
| LVEF | -0.13 | <0.001 | -0.13 | <0.001 | -0.18 | <0.001 | 0.78 | -0.14 | <0.001 | -0.12 | <0.001 | 0.33 |
| LVEDV index | 0.22 | <0.001 | 0.2 | <0.001 | 0.24 | <0.001 | 0.76 | 0.24 | <0.001 | 0.20 | <0.001 | 0.01 |
| LV mass index | 0.21 | <0.001 | 0.25 | <0.001 | 0.16 | <0.001 | <0.001 | 0.22 | <0.001 | 0.23 | <0.001 | 0.012 |

Model adjusted for age, race, gender, smoking status, body mass index, prevalent hypertension, prevalent diabetes, systolic blood pressure, heart rate, serum fasting glucose.
Abbreviations- LV: left ventricular, LVEF: left ventricular ejection fraction, LVEDV: left ventricular diastolic volume

Specifically, there was an interaction between sex and QRS duration, such that longer QRS duration in men was more strongly associated with higher LV mass (P-int=0.012) and higher LVEDV index (P-int=0.01) as compared to women. Furthermore, an interaction was observed between race and QRS duration, such that longer QRS duration was more strongly associated with LV mass index in Black participants as compared to White participants (P-int<0.001) (**Table 2**).

The cumulative incidence of primary outcome events stratified by QRS interval quartiles are shown in **Figure 3**. Participants in the highest quartile of QRS duration experienced 110 primary outcome events and had the highest cumulative incidence rate of MACE (log-rank P=0.026). In Cox regression analysis, longer QRS duration was associated with higher risk of MACE (HR = 4.38 [95% CI: 2.12, 9.06]). However, after adjustment for CV risk factors, this association attenuated and was no longer significant (HR = 1.85 [95% CI: 0.9, 3.79]) (**Table 3**).

**Figure 3:**
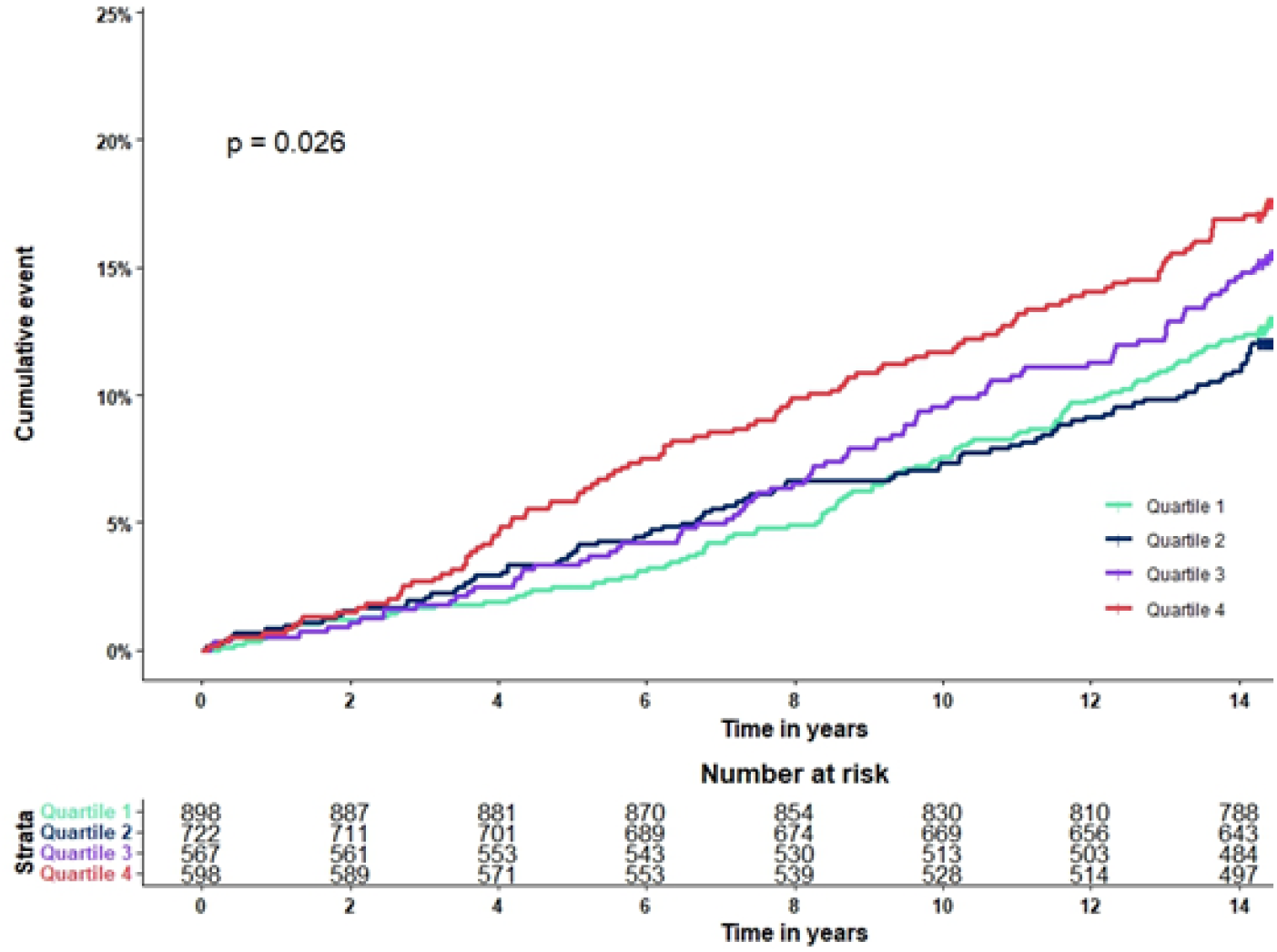
Cumulative Incidence of Major Adverse Cardiac Events Across Quartiles of QRS Duration Cumulative incidence rate across quartiles of QRS duration compared using log-rank testing.

**Table 3:** Association of log transformed QRS duration with risk of major adverse cardiac events

|  |  | Unadjusted |  |  | Adjusted |  |  |
| --- | --- | --- | --- | --- | --- | --- | --- |
|  | Events (N) | HR (95% CI) | p-value | p-int* | HR (95% CI) | p-value | p-int* |
| Total | 423 | 4.38(2.12,9.06) | <0.001 | - | 1.85(0.90,3.79) | 0.10 | - |
| Black | 274 | 7.31(3.14,17.0) | <0.001 | 0.4 | 1.91(0.80,4.55) | 0.14 | 0.8 |
| White | 106 | 3.23(0.64,16.2) | 0.2 |  | 1.19(0.23,6.21) | 0.8 |  |
| Men | 230 | 1.00(0.3 x1,3.20) | >0.9 | 0.023 | 1.33(0.46,3.87) | 0.6 | 0.3 |
| Women | 193 | 6.66(2.32,19.1) | <0.001 |  | 2.45(0.94,6.39) | 0.067 |  |

Adjusted for QRS, age, race, gender, smoking status, body mass index, prevalent hypertension, prevalent diabetes, systolic blood pressure, heart rate, serum fasting glucose.
Race was not included as an adjustment in the race-based subgroups
Sex was not included as an adjustment in the sex-based subgroups
Interaction testing was performed for Black race\*QRS duration and sex\*QRS duration

In subgroup analysis, QRS duration was not associated with risk of MACE in White participants or men. However, longer QRS duration was significantly associated with risk of MACE in women (HR = 6.66 [95% CI: 2.32, 19.1). This association was attenuated after adjustment for CV risk factors with a trend toward significance (HR = 2.45 [95% CI: 0.94, 6.39]). Similarly, prolonged QRS duration was associated with higher risk of MACE among Black participants (HR = 7.31 [95% CI: 3.14, 17]) in the unadjusted analysis, but this association did not remain significant in the adjusted model **Table 3**. No significant interaction was observed between sex and QRS duration or race and QRS duration for risk of the primary outcome.

## Discussion

In this multiethnic cohort of adults without CVD, we made several observations. First, prolonged QRS interval was associated with greater LV hypertrophy and worse LV systolic function independent of CV risk factors. Second, prolonged QRS duration was more steeply associated with LV hypertrophy in men as compared to women and in Black participants as compared to White participants. Finally, longer QRS duration was associated with greater risk of MACE, which appeared to be mediated by prevalent CV risk factors. Among women with prolonged QRS duration, there was a non-significant trend toward excess risk of MACE, independent of CV risk factors. We found that the prognostic implications of prolonged QRS duration in healthy adults did not differ significantly by race or sex.

Prior studies examining the association of QRS duration with risk of adverse outcomes have yielded varied results. An analysis of the Framingham cohort showed that QRS duration was not significantly associated with risk of CVD, CV death, or sudden cardiac death(25). This pattern of association was consistent in men and women, though women had a non-significant trend towards higher relative risk of adverse outcomes compared to men. In a study of over 10,000 healthy adults (51% men) from Finland, investigators found that QRS duration > 110 ms was associated with greater risk of all cause death, CV mortality, and sudden cardiac death(14). This association was independent of CV risk factors and has been replicated in subsequent studies(15, 26). Notably, these study populations consisted of mostly White participants. Although we did not observe an association between QRS duration and risk of MACE after adjustment for CV risk factors, there was a trend toward significance, which could be attributed to the smaller sample size and lower event rate in our study. Mechanistic analysis showed that QRS prolongation was significantly associated with LV hypertrophy and reduced LV systolic function in healthy adults, a finding that is in keeping with what has previously been reported in the literature(27, 28).

The present study showed that there are differences in QRS duration at baseline in men vs women and Black vs White adults without CVD. Our findings are consistent with prior studies, which have reported lower QRS duration among women and Black individuals(29, 30, 31). Shorter QRS duration in women has been attributed to lower LV mass in women as compared to men(32). The mechanism driving shorter QRS duration in Black adults is not well understood. It is noteworthy that while Black participants had shorter QRS duration compared to White participants, longer QRS duration was more steeply associated with higher LV mass among Black participants, a finding that to our knowledge has not been previously reported.

Limited data is available regarding the prognostic implications of QRS duration in healthy adults across sex- and race-based subgroups. In an analysis of The Strong Heart Study cohort, investigators evaluated the association of QRS duration with risk of CVD in American Indians. Similar to what we observed, prolonged QRS duration in American Indian women but not men, was associated with greater risk of CVD(33). Although women with longer QRS duration numerically had higher risk of MACE compared to men, no significant interaction was noted. A similar pattern of association in Black vs White participants. Taken together, the present study suggests there are no significant differences in the prognostic implications of prolonged QRS duration in race- or sex-based subgroups. More studies are needed to confirm this finding.

CV risk factors and LV mass both have been implicated in the development of prolonged QRS duration(31). In women, it has been suggested that differences in hormone levels may affect gap junction expression such that women are more resistant to conduction abnormalities(30, 35). Shorter QRS duration in Black individuals has been previously reported but is poorly understood. Likely genetic differences in myocardial conduction underlie differences in QRS duration across ethnicities, as ethnic heterogeneity in SNPs implicated in QT prolongation have been reported(36). More studies are needed to understand the mechanisms driving differences in QRS duration among ethnic groups. Furthermore, additional investigation is needed to understand why QRS prolongation reflections greater LV hypertrophy in Blacks compared to Whites, despite Blacks having shorter QRS duration.

These findings have clinical implications. The QRS interval is a readily available clinical data point that can offer significant prognostic information. Accordingly, the 2018 ACC/AHA/HRS guideline on the evaluation and management of patients with bradycardia and cardiac conduction delay recommend checking a transthoracic echocardiogram in healthy adults with a LBBB(19). Our findings suggest that prolonged QRS duration, regardless of the presence of LBBB, may warrant assessment of LV structure and function, particularly in Black men. Furthermore, uniform QRS interval cut-offs are used to guide clinical decision making, specifically with regard to CRT. Our study suggests caution in using uniform QRS duration cut offs across different demographic groups to inform management strategies.

Our study does have limitations that merit mention. First, the outcome of sudden cardiac death was not available in the DHS, an adverse event known to be associated with QRS interval prolongation. Second, owing to the small number of participants with RBBB, LBBB, or NIVCD, association of QRS duration with adverse events in participants stratified by QRS morphology could not be evaluated. Finally, given the study was observational, it remains susceptible to residual confounding.

In conclusion, prolonged QRS duration is associated with higher risk of MACE in healthy adults, primarily mediated by prevalent CV risk factors. Risk of adverse of events attributed to QRS prolongation is consistent across race- and sex-based subgroups. However, QRS duration and the relationship of QRS prolongation to underlying LV hypertrophy vary by sex and race. Our findings suggest caution should be taken in using uniform QRS cut offs to inform clinically decision making.

## Data Availability

The data referred to in the manuscript is available by request to the Dallas Heart Study

